# Shared proteomic landscape between arteriosclerosis and cardiovascular endpoints: a Mendelian randomization and observational study integrating AlphaFold3 for structural prediction

**DOI:** 10.1101/2025.05.15.25327712

**Authors:** Jingxian Huang, Margaux Achtari, Devendra Meena, Alexander Smith, Renae Judy, Nour Mimouni, Joshua C. Bis, Nora Franceschini, Scott M. Damrauer, Ioanna Tzoulaki, Abbas Dehghan

**Author notes:** Corresponding authors (A. Dehghan); (D. Meena). Joint first author (Jingxian Huang, Margaux Achtari, Devendra Meena).

## Abstract

**Background:** Atherosclerosis and arteriosclerosis are major contributors to cardiovascular disease (CVD), yet their shared and distinct molecular underpinnings remain incompletely understood. This study integrates proteomics, Bayesian colocalization, and Mendelian randomization (MR), and structural modelling to explore the shared and distinct plasma proteome associated with arteriosclerosis and atherosclerosis across different vascular beds.

**Methods:** We leveraged *cis*-pQTLs for 5,813 unique proteins from the UK Biobank (UKB) Pharma Proteomics Project (N=54,219) and deCODE genetics (N=35,559) and assessed the association with five arteriosclerotic/atherosclerotic markers, as well as eight cardiovascular events, using Bayesian colocalization and bidirectional MR. We validated the findings through tissue-specific transcriptomics, observational data from UKB, and AlphaFold3 for structural prediction. Finally, mediation analysis evaluated the role of vascular traits in linking proteins to CVD risk.

**Results:** We prioritized ten proteins potentially causally associated with both the arteriosclerotic/atherosclerotic markers and cardiovascular events. Five of them (ANGPTL4, APOB, BRAP, LPA, and ZPR1), were associated with increased levels of arteriosclerosis/ atherosclerosis and risk of CVD, whereas four (DUSP13, FN1, IL6R, and MMP12) were associated with reduced levels of arteriosclerosis/ atherosclerosis and risk of CVD. ABO was associated with increased risk of peripheral artery disease (PAD) and CVD but inversely related to ASI. Mediation analyses estimated that LPA’s effect on stroke was primarily mediated through carotid plaque (92.4%), while DUSP13’s effect on coronary artery disease was primarily mediated via PAD (91.0%). Observational analyses and transcriptomic validation corroborated these associations. Structural modelling using AlphaFold3 identified key functional variants in several proteins, including ANGPTL4 and FN1, potentially underlying the pathogenic mechanists.

**Conclusions:** The present study elucidates the shared and distinct proteomic signatures across arteriosclerosis, atherosclerosis, and cardiovascular disease, underscoring the importance of vascular-bed-specific mechanisms. These identified proteins offer promising avenues for biomarker-driven risk stratification and therapeutic interventions, with potential for dual-purpose interventions across vascular territories.

## Introduction

Atherosclerotic cardiovascular disease (ASCVD) remains the leading cause of mortality worldwide, highlighting the urgent need for improved preventive strategies and therapeutic targets ^1^. Arteriosclerosis (the hardening and loss of elasticity of the arteries) and atherosclerosis (the deposition of lipids on the arterial intima) are the key pathological processes underlying CVDs in various vascular beds, including coronary artery disease (CAD) in coronary arteries, stroke in carotid and cerebral arteries, and peripheral artery disease (PAD) in the arteries of the limbs. Subclinical markers of atherosclerosis and arteriosclerosis provide valuable opportunities to investigate these two pathologies before clinical events arise. Arterial stiffness index (ASI), coronary artery calcification (CAC), carotid plaque, carotid intima-media thickness (cIMT) and PAD each reflect different aspects of vascular remodelling and disease progression in various vascular beds ^2–5^. Increasing evidence indicates common molecular pathways and pathological drivers between these conditions, including chronic inflammation, oxidative stress, and endothelial dysfunction ^6,7^. While these markers are associated with future cardiovascular events and show certain common pathways, they may differ in their predictive ability. For instance, carotid plaque may serve as a stronger predictor of atherosclerotic clinical events than cIMT ^8^. Moreover, there might be other underlying pathways specific to one of these pathologies.

Circulating proteins are thought to play various roles in metabolic, signalling, and physiological processes and are key biomarkers of the progression of atherosclerosis ^9,10^. The plasma proteome reflects contributions from nearly all tissues, with proteins entering the bloodstream via active secretion, passive diffusion, or cellular turnover ^11^. Importantly, these proteins often mirror pathological processes occurring in tissues such as the endothelium, liver, or vasculature, making blood an accessible window into otherwise inaccessible sites. By leveraging genetic instruments and integrative analyses, circulating proteins can reveal causal mechanisms across tissues, offering insights into shared pathophysiology and guiding translational efforts in cardiovascular disease. Recent large-scale proteogenomic studies have identified thousands of protein quantitative trait loci (pQTLs) associated with protein abundance providing valuable resources for causal inference ^12,13^. Mendelian randomization (MR) and Bayesian colocalization analyses are utilized to leverage genetic variants as instrumental variables (IVs), providing stronger evidence for causal relationships and evaluating shared causal variants between protein levels and the outcomes of interest ^14,15^.

This study aims to highlight the commonality and differences in circulating proteins associated with arteriosclerosis and atherosclerosis within different vascular beds. Moreover, the arteriosclerosis- and atherosclerosis-associated proteins were further evaluated through downstream analysis evaluating their association with cardiovascular events to prioritize a set of proteins that shared between the two aspects. This approach aims to provide insights into how arteriosclerosis and atherosclerosis may manifest similarly or differently across vascular territories and how they are associated with cardiovascular events. Finally, this study emphasizes triangulating findings through observational analyses and causal inferences within disease-relevant tissues, while identifying potential therapeutic targets that exhibit joint effects on subclinical atherosclerosis and CVD. The novelty of this study lies in its use of an integrated approach to combine observational and Mendelian randomisation approaches to prioritise potential causal proteins and the use of Alphafold3 for structural prediction. This will provide novel insights into how these pathological processes manifest across vascular territories, potentially suggesting vascular-bed-specific therapeutic targets with combined effects on subclinical atherosclerosis and cardiovascular disease.

## Methods

We first performed colocalization to identify shared genetic signals between plasma proteins and arteriosclerotic/atherosclerotic markers. We then applied MR for causal inference, followed by transcriptomic validation, observational analyses, mediation modelling, and 3D structural predictions using AlphaFold3 (**Figure 1**).

**Figure 1.**
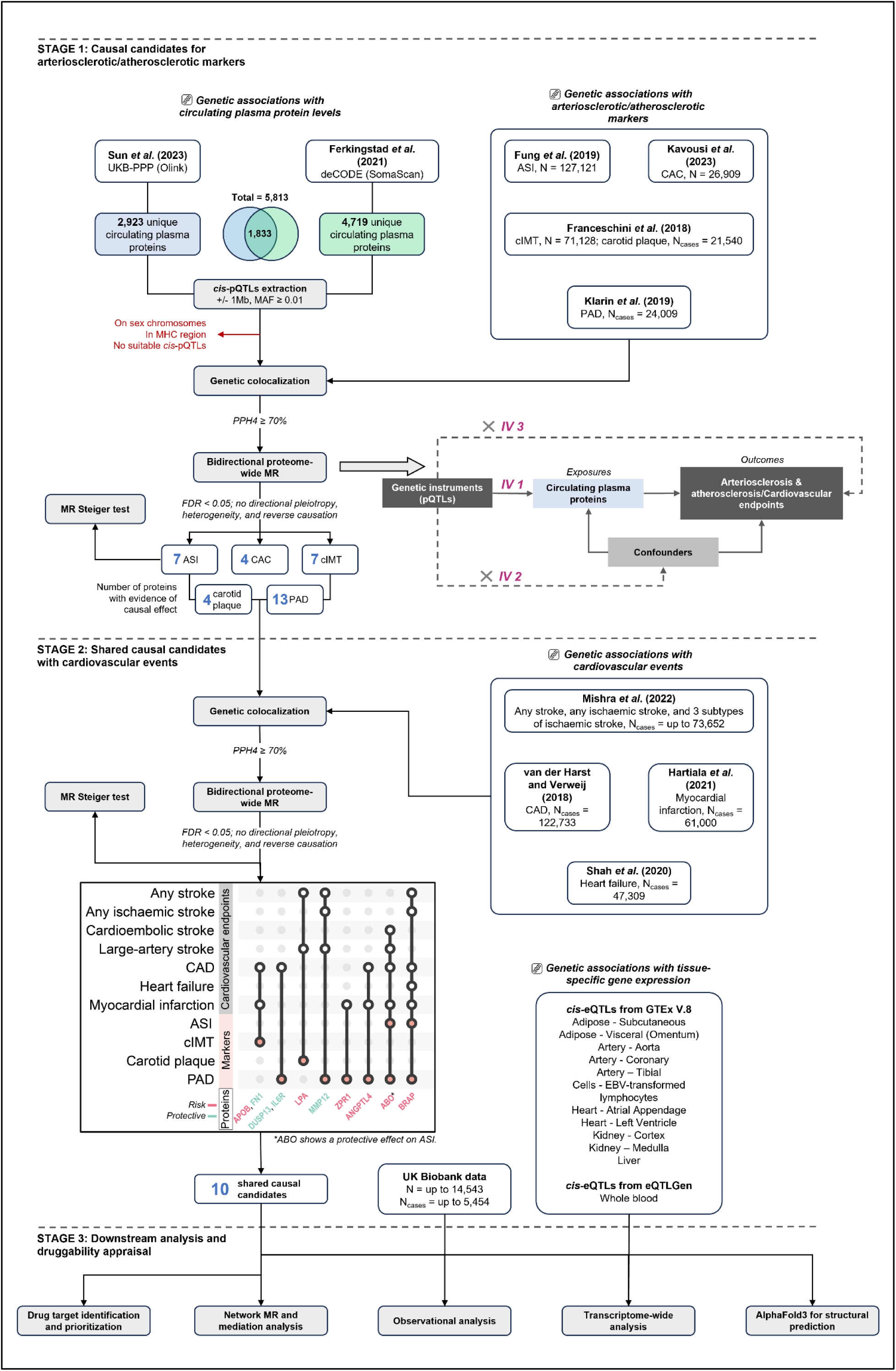
A flowchart depicting the study design and a schematic representation of the cis-MR approach. ASI, arterial stiffness index; CAC, coronary artery calcification; cIMT, carotid artery intima-media thickness; PAD, peripheral artery disease; pQTLs, protein quantitative trait loci; MR, Mendelian randomization; CAD, coronary artery disease; eQTLs, expression quantitative trait loci.

### GWAS for plasma protein levels

GWAS summary statistics for up to 5,813 unique circulating plasma proteins were obtained from publicly available data published by the UK Biobank Pharma Proteomics Project (UKB-PPP) ^16^ and deCODE genetics ^17^. The UKB-PPP studied the proteomic profiles across 54,219 UKB participants, of predominantly European ancestry, whose blood plasma samples were measured using the antibody-based Olink Explore 3072 proximity extension assay (PEA), capturing 2,923 unique proteins. deCODE genetics measured levels of 4,719 plasma proteins in 35,559 Icelanders using the aptamer-based SomaScan multiplex aptamer assay (version 4). 31.5% of the unique proteins (1,833 out of 5,813) were available in both study (**Figure 1**).

### GWAS for markers of arteriosclerosis and atherosclerosis

Five different arteriosclerotic and/or atherosclerotic markers were investigated in this study. Genetic associations for ASI were obtained from a GWAS of 127,121 UK Biobank (UKB) individuals of European ancestry ^2^. ASI was derived using a pulse waveform which was measured at the finger for 10–15 seconds. Genetic associations for CAC were obtained from the largest GWAS meta-analysis across participants from the Cohorts for Heart and Aging Research in Genomic Epidemiology (CHARGE) consortium and collaborators, comprising 26,909 individuals of European ancestry from 16 cohorts ^4^. CAC was measured by computed tomography. GWAS summary statistics for cIMT and carotid plaque were obtained from a meta-analysis of GWAS in European ancestral individuals from the CHARGE consortium and the University College London-Edinburgh-Bristol (UCLEB) consortium, comprising up to 71,128 participants for cIMT and up to 48,434 participants for carotid plaque (N_cases_ = 21,540) ^3^. Both measures were evaluated employing high-resolution ultrasonography, and the carotid plaque was defined by atherosclerotic thickening of the common carotid artery wall or by a surrogate measure indicating more than 25% narrowing of the lumen. Finally, genetic veterans of European ancestry in the United States (N_cases_ = 24,009) ^5^. ASI, CAC, and cIMT were considered as continuous traits while carotid plaque and PAD were considered as dichotomous traits.

### GWAS for cardiovascular events

We obtained summary statistics for any stroke (N_cases_ = 73,652), any ischaemic stroke (N_cases_ = 62,100), and subtypes of ischaemic stroke (including large-artery stroke, N_cases_ = 6,399; cardioembolic stroke, N_cases_ = 10,804; and small-vessel stroke, N_cases_ = 6,811) from the GIGASTROKE consortium ^18^. Summary statistics for coronary artery disease (CAD) were obtained from the UK Biobank CARDIoGRAMplusC4D meta-analysis collectively with 122,733 cases and 547,261 controls ^19^. GWAS summary statistics for myocardial infarction were obtained from a meta-analysis of the UKB and CARDIoGRAMplusC4D consortium with ∼61,000 cases and ∼578,000 controls ^20^. Last, genetic associations for heart failure were obtained from a GWAS meta-analysis across 26 studies of the Heart Failure Molecular Epidemiology for Therapeutic Targets (HERMES) Consortium, comprising 47,309 cases and 930,014 controls ^21^.

### Bayesian colocalization analysis

We performed genetic colocalization analysis using a Bayesian framework to investigate if plasma proteins and outcomes of interest (i.e., arteriosclerotic/atherosclerosis markers and cardiovascular events) share the same causal variant ^22^. This approach could determine if any potential association is due to the pleiotropic effects of variants in linkage disequilibrium (LD) ^23^. Using the “coloc” package in R ^22^, we performed pairwise colocalization on the *cis*-region of each gene coding the proteins, which was defined as the 1 megabase (Mb) region from both sides of the coding region. Rare (minor allele frequency (MAF) < 0.01) and non-biallelic variants were excluded prior to the analysis. The prior probability that a genetic variant in the pre-defined region is exclusively associated with one of the two traits was set to 1 × 10^−5^ and the prior probability associated with both traits was set to 1 × 10^−5^. Each configuration of the two traits could be assigned to one of the hypotheses:

- H0: no association with either the protein or outcome of interest.
- H1: association with the protein but not outcome of interest.
- H2: association with outcome of interest but not the protein.
- H3: association with both the protein and outcome of interest, but at separate causal variants.
- H4: association with the protein and outcome of interest at a shared causal variant.

Proteins encoded by genes located on the sex chromosomes were excluded from the colocalization analysis. Additionally, due to the complex LD structure of genetic variants in the human Major Histocompatibility Complex (MHC) region, proteins encoded by genes within this region were also excluded from genetic colocalization and all downstream analyses. We considered the posterior probability (PP) of H4 ≥ 70% as strong evidence of colocalization. Protein-trait associations were dropped for further analysis if they did not reach the threshold (**Figure 1**).

### Proteome-wide Mendelian randomization analysis

We performed two-sample univariable MR to investigate the potential causal relationship between the colocalization prioritised circulating levels of plasma proteins and outcomes of interest, using *cis*-acting protein quantitative trait loci (*cis*-pQTLs) as IVs. Valid MR estimates rely on three key assumptions, including (1) relevance, (2) independence, and (3) exclusion restriction. We extracted biallelic SNPs located within ± 1MB of the gene encoding the protein (i.e., *cis*-pQTLs), and further filtered on genome-wide significance level (P < 5 × 10^−8^), MAF ≥ 0.01, and instrumental strength (F-statistics ≥ 10). The protein dataset was then harmonized against the outcome dataset employing the “harmonise_data” function from the “TwoSampleMR” R package ^24^ clumped at a 10,000 kilobase (Kb) window with an LD threshold of r^2^ < 0.001 employing the “ld_clump” function from the “ieugwasr” R package ^25^ to obtain independent IVs. The Wald ratio was used as the primary MR method when the number of IVs was fewer than 2, while the inverse variance weighted (IVW) method was employed when≥2 IVs were available. Sensitivity analyses were conducted using the weighted median (WM) and MR-Egger methods when ≥3 IVs were available. Cochran’s Q test was performed to assess potential heterogeneity presented among the selected IVs. MR analysis was performed bidirectionally to assess potential reverse causation. We applied Benjamini-Hochberg false discovery rate (FDR) correction to control for multiple testing, and an FDR-corrected P-value < 0.05 was considered statistically significant. All MR results were evaluated based on FDR-corrected P-value < 0.05, no evidence of heterogeneity (Cochran’s Q P-value ≥ 0.05), no evidence of horizontal pleiotropy (P_Egger intercept_ ≥ 0.05), and no evidence of reverse causality. Protein-trait associations pass above filters were prioritized for further downstream analysis. MR Steiger was further conducted on these associations as an additional sensitivity check to orient the direction of causality ^26^.

### Utilizing tissue-specific transcriptomic data with SMR and HEIDI

We conducted summary data-based MR (SMR) and heterogeneity in dependent instruments (HEIDI) tests to integrate GWAS and expression quantitative trait loci (eQTLs) data to identify potential causal genes whose expression levels are linked to arteriosclerotic/atherosclerotic markers and cardiovascular events ^27^. The SMR and HEIDI methodology can be interpreted as an data from eQTLGen (n = 31,684) for whole blood and GTEx v.8 ^28^ for 12 tissues relevant to the CVD pathology. This included adipose-subcutaneous, adipose-visceral (omentum), artery-aorta, artery-coronary, artery-tibial, cells-ebv-transformed lymphocytes, heart-atrial appendage, heart-left ventricle, kidney-cortex, kidney-medulla, and liver (**Figure 1**). The eQTL dataset was publicly available at the eQTLGen consortium ^29^. Version 8 release of the GTEx eQTL summary data can be downloaded from https://yanglab.westlake.edu.cn/software/smr/#DataResource. The default p-value threshold to select the top associated eQTL for the SMR test is 5 × 10^−8^. By default, we only run the SMR analysis in the cis regions. The default threshold of the eQTL P-value to select eQTLs for the HEIDI test is 1.5 × 10^−3^. All other settings were left as default. We performed FDR correction to account for multiple testing burdens (FDR> 0.05).

### Triangulation with observational analysis

We further conducted an observational analysis on the association of circulatory proteins measured by OLINK in UKB with measures on atherosclerosis and arteriosclerosis, collected at the point of blood sample collection. This included cIMT (mean of UKB fields 22672, 22675, 22678 and 22681) and ASI (Field 21021). Incident cases of PAD, heart failure, CAD, myocardial infarction, all stroke and ischaemic stroke were identified in the latest release of patient-linked GP records, Hospital Episode Statistics (HES) and death registry using the same ICD-10 code lists for each condition as their respective GWAS study. Follow-up time was calculated from the date of blood collection for each individual to the occurrence of the disease, death, or loss to follow-up, or censoring time, whichever came first. We did not include ischaemic stroke subtypes, CAC, and carotid plaque as they were assessed and identified using imaging-based phenotypes. Each protein level was rank-inverse normalised and adjusted for sex and age at the time of blood collection to match their respective GWAS procedures. We standardised the residuals of the protein levels and model covariates using rank-inverse normalisation and used the standardised values for association testing using linear regression (ASI, cIMT) and Cox proportional hazard models (PAD, heart failure, CAD, myocardial infarction, all stroke and ischaemic stroke) adjusted for age at blood sample collection, sex, sample age, ethnicity, BMI, Townsend deprivation index, the first 20 genetic principle components, UKB assessment centre, UKB array type, OLINK batch and UKB-PPP subcohort. P-values were adjusted for 5% FDR across all association tests.

### Network MR and mediation analysis

We conducted network MR and mediation analysis for proteins causally associated with both arteriosclerotic/atherosclerotic markers and cardiovascular events to investigate the effects of proteins (exposure) on cardiovascular events (outcome) operating through arteriosclerotic/atherosclerotic markers (mediator). Specifically, we obtained the effects of proteins on cardiovascular events (total effect; β_EO_) and the effects of proteins on arteriosclerotic/atherosclerotic markers (β_EM_) from the primary proteome-wide MR analysis. We then performed univariable MR to estimate the effects of arteriosclerotic/atherosclerotic markers on cardiovascular events (β_MO_). To identify and exclude potential IVs that may violate the assumptions of MR, we utilized the Mendelian Randomization Pleiotropy Residual Sum and Outlier test (MR-PRESSO) in this step ^30^. The potential causal effects of cardiovascular events on arteriosclerotic/atherosclerotic markers were also assessed to orientate potential bidirectional effects. Last, we decomposed the indirect effects following the product method by calculating the product of β_EM_ and β_MO_^31^. To quantify the pathway-specific contributions to the total effects, we calculated the proportion mediated by subtracting the indirect effects from β_EO_. Standard error was estimated using the delta method ^32^.

### 3D structure prediction with AlphaFold3

We assessed whether the pQTLs used as IVs in the proteome-wide MR analysis were either missense variants themselves or in strong LD (*r*^2^ > 0.8) with a missense variant affecting the corresponding protein. We then integrated AlphaFold3 (https://alphafoldserver.com/) ^33^ to predict the 3D structural alterations resulting from the missense variant and visualized them in the PyMol software (https://pymol.org).

### Investigation of approved and investigational drugs

We queried the proteins shared between arteriosclerotic/atherosclerotic markers and cardiovascular events to the Open Targets Known Drugs database (https://platform.opentargets.org/) to retrieve drug, disease, and clinical trial information. We additionally cross-referenced these proteins to the DrugBank database Release Version 5.1.13 (https://go.drugbank.com/releases/latest) to retrieve approved drug information and detailed indications.

## Results

### Potential causal proteins prioritised for arteriosclerotic/atherosclerotic markers

We performed colocalisation for up to 3,560 unique proteins (2,796 from UKB-PPP and 1,745 from deCODE; of these, 981 were available in both) with suitable *cis*-variants at an intercepted genomic region with the 5 arteriosclerotic/atherosclerotic makers, representing 17,181 unique pairs (protein-trait). A total of 62 unique proteins showed strong evidence of colocalisation (PP.H4 ≥ 70% in UKB-PPP or deCODE; **ST1-2**) with at least one arteriosclerosis/atherosclerosis marker. Specifically, we identified 10 proteins for ASI (of 3,560 tested), 10 for CAC (of 3,188), 9 for cIMT (of 3,560), 11 for carotid plaque (of 3,560), and 29 for PAD (of 3,313) (**Figure 2A-B**). Among the 62 proteins, 20 were available in both panels, with 9 demonstrating evidence of colocalization in both (**Figure 2B**).

**Figure 2.**
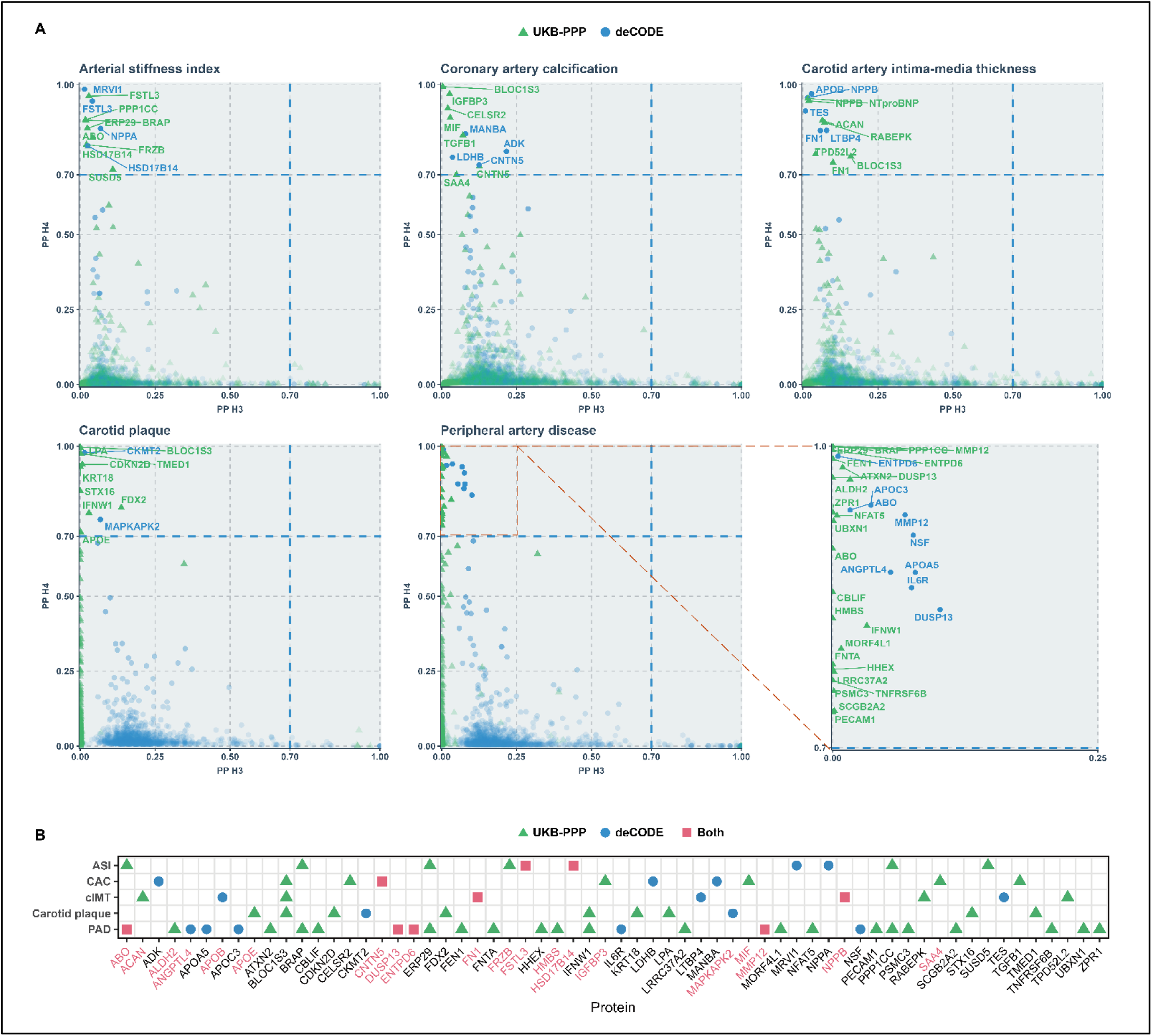
Genetic colocalization for arteriosclerotic/atherosclerotic markers and plasma proteins. **A.** Scatter plot showing PPH4 against PPH3 for each protein from Bayesian genetic colocalization analysis. PPH4 is the posterior probability of shared causal variants; PPH3 is the posterior probability of distinct causal variants. Plasma proteins with strong evidence of colocalization (PPH4 ≥ 70%) were highlighted and labelled. **B.** Heatmap showing proteins with evidence of colocalization across 5 arteriosclerotic/atherosclerotic markers. The point shape represents the proteomics panel from which the colocalized proteins originate. Proteins available and tested in both panels are highlighted in red on the x-axis.

We subsequently conducted MR analysis targeting protein-trait associations with strong evidence of colocalization (PPH4 ≥ 70%). After applying rigorous filters for statistical significance, horizontal pleiotropy, heterogeneity, and potential reverse causality, we identified and prioritized 7 proteins for ASI, 4 for CAC, 7 for cIMT, 4 for carotid plaque, and 13 for PAD (**Figure 3; ST3**). 33.3% of the protein candidates were assessed using three or more IVs and were further studied for the potential pleiotropic effect of the instruments using at least one sensitivity MR method. The five arteriosclerotic/atherosclerotic markers exhibited no overlap in protein candidates, except for the histo-blood group ABO system transferase (ABO) and the BRCA1-associated protein (BRAP), which were shared between ASI and PAD. Notably, genetically proxied ABO levels showed an inverse association with ASI (Beta: −0.02, 95% confidence interval (CI): −0.03 to −0.01) but were positively associated with PAD risk (Odds ratio (OR): 1.07, 95% CI: 1.03 to 1.12). The association of HSD17B14 with ASI disappeared after the removal of invalid IVs identified by MR Steiger test, indicating potential reverse causality (**ST3**).

**Figure 3.**
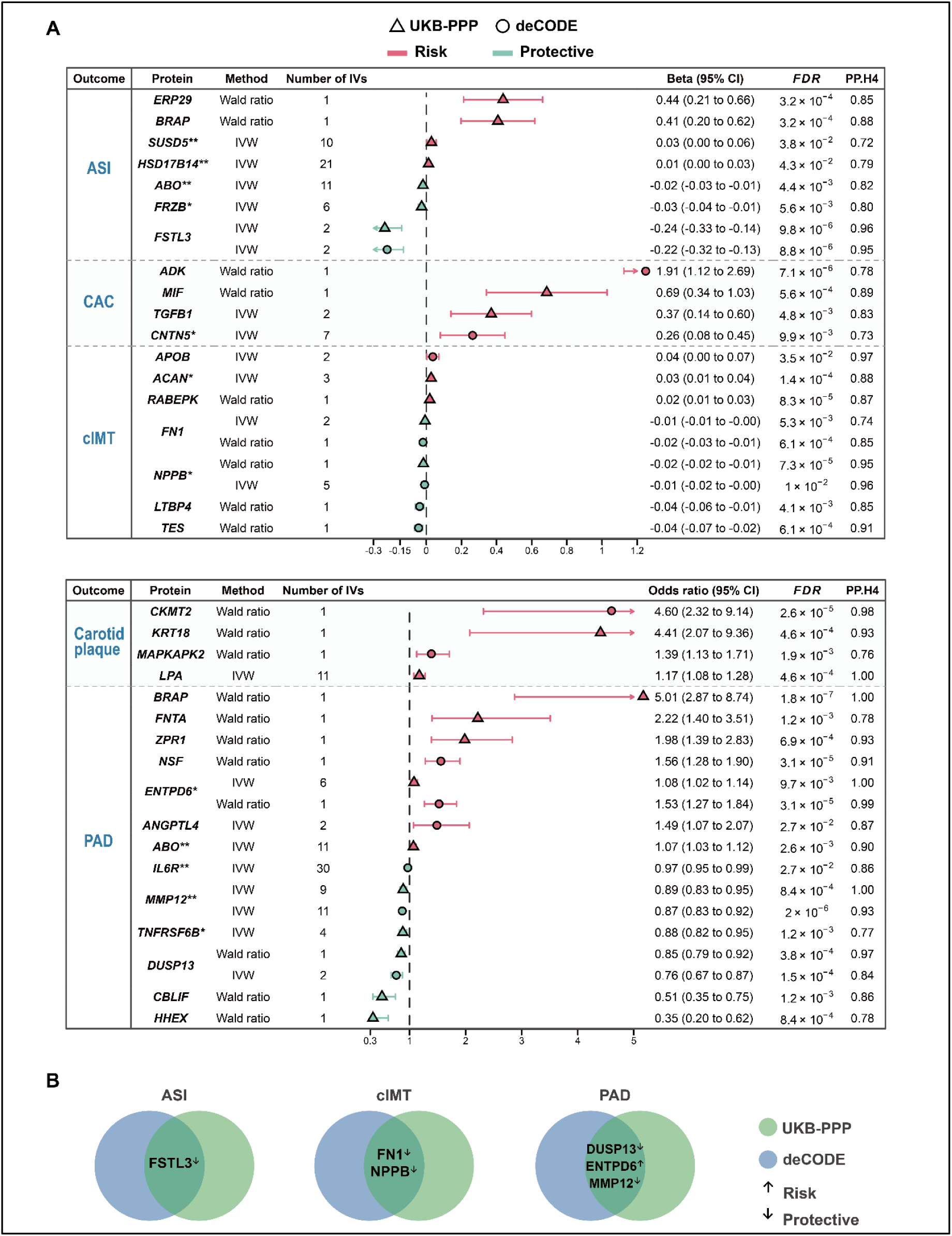
Prioritized protein candidates for arteriosclerotic/atherosclerotic markers in Mendelian randomization analysis. **A.** MR causal estimates of prioritized circulating protein levels with arteriosclerotic/atherosclerotic markers from UKB-PPP and deCODE. The upper panel shows continuous outcomes while the lower panel shows dichotomous outcomes; The shape of the estimate points indicates the study source; All estimates correspond to a one-standard-deviation increase in protein levels; *, supported by one of the sensitivity MR methods (WM or MR-Egger); **, supported by both the sensitivity MR methods (WM and MR-Egger). **B.** Candidates prioritized by both UKB-PPP and deCODE. ASI, arterial stiffness index; CAC, coronary artery calcification; cIMT, carotid artery intima-media thickness; PAD, peripheral artery disease; IVs, instrumental variables.

### Genetic colocalization and MR analysis with cardiovascular events

We took forward 33 proteins associated with at least one arteriosclerotic/atherosclerotic marker for further investigation by firstly performing genetic colocalization analyses with eight cardiovascular events. Protein-trait associations with strong colocalization evidence (PPH4 ≥ 70%) were then assessed for potential causal relationships using MR analysis. After applying the same rigorous filters for causal appraisal, we prioritized 11 protein candidates that were shared between arteriosclerotic/atherosclerotic markers and cardiovascular events, combining evidence from both UKB-PPP and deCODE (**Figure 4A; ST4**). The MR Steiger test did not identify any invalid IV among these associations (**ST4**).

**Figure 4.**
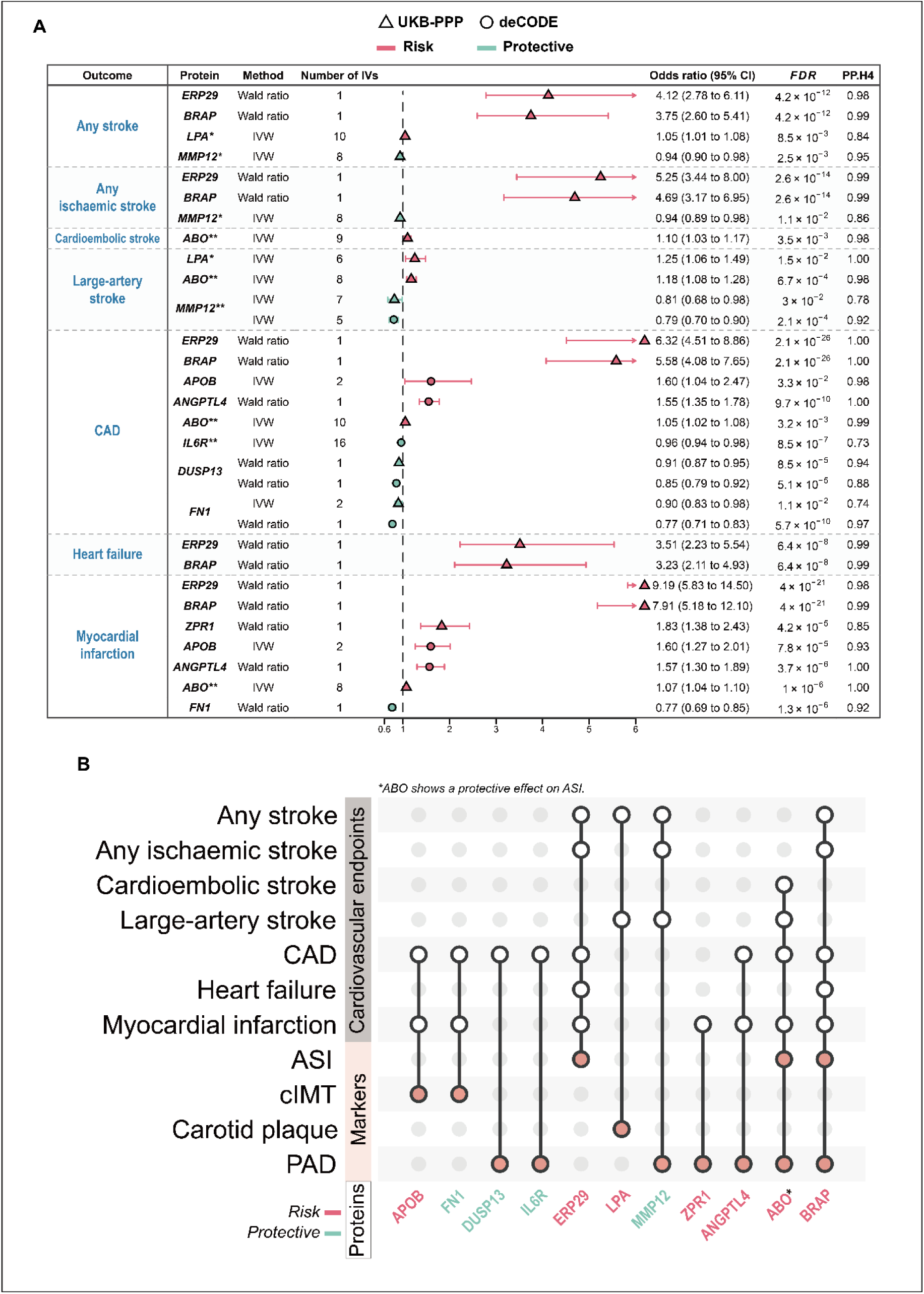
Prioritized protein candidates for arteriosclerotic/atherosclerotic markers that were shared with cardiovascular events. **A.** MR causal estimates of shared candidates on cardiovascular events from UKB-PPP and deCODE. The shape of the estimate points indicates the study source; All estimates correspond to a one-standard-deviation increase in protein levels; *, supported by one of the sensitivity MR methods (WM or MR-Egger); **, supported by both the sensitivity MR methods (WM and MR-Egger). **B.** UpSet plot summarizing overlaps of potential causal proteins between cardiovascular events and arteriosclerotic/atherosclerotic markers. ASI, arterial stiffness index; CAC, coronary artery calcification; cIMT, carotid artery intima-media thickness; PAD, peripheral artery disease; CAD, coronary artery disease; IVs, instrumental variables.

Among the 11 proteins identified, endoplasmic reticulum protein 29 (ERP29) and BRAP emerged as the most pleiotropic candidates, being associated with five distinct cardiovascular events (**Figure 4B**). PAD-associated proteins were the most pleiotropic, being associated with multiple other cardiovascular events, particularly with CAD (**Figure 4B**). For example, genetically determined levels of the dual specificity protein phosphatase 13A (DUSP13) and the interleukin-6 receptor subunit alpha (IL6R) were associated with a lower risk of PAD (OR: 0.85 per-SD higher DUSP13; OR: 0.97 per-SD higher IL6R) and CAD (OR: 0.91 per-SD higher DUSP13; OR: 0.96 per-SD higher IL6R); genetically determined levels of the angiopoietin-related protein 4 (ANGPTL4) and ABO were associated with a higher risk of PAD (OR: 1.49 per-SD higher ANGPTL4; OR: 1.07 per-SD higher ABO) and CAD (OR: 1.55 per-SD higher ANGPTL4; OR: 1.05 per-SD higher ABO). Notably, we found no overlap of causal candidates between CAC and cardiovascular events.

### Sensitivity test in gene-dense regions

Among the 11 protein candidates we highlighted, BRAP and ERP29 are encoded by genes located in close genomic proximity on chromosome 12. We re-conducted colocalisation restricting to only the coding region of these two proteins to minimise confounding by including regulatory variants from neighbouring genes. Colocalisation results indicated that ERP29 no longer shares the same causal variant with ASI and PAD (PPH4 < 1%), while the probability of BRAP sharing the same causal variant with ASI was significantly attenuated (PPH4 < 20%). Nonetheless, BRAP still exhibited strong evidence of colocalisation with PAD, and further with any stroke, ischemic stroke, CAD, myocardial infarction, and heart failure (PPH4 > 98%; **ST5**). These findings suggested that BRAP is the more likely causal candidate in this genomic region, whereas ERP29 associations may have been driven by LD rather than a shared causal variant. Therefore, we removed ERP29 from any downstream analysis.

### SMR and HEIDI analysis validated the association of *ABO* expression with ASI, PAD, and cardiovascular events

We subsequently performed SMR and HEIDI analysis on the 10 highlighted proteins and their associated traits to assess the causal relationships between gene expression levels and outcomes of interest in disease-relevant tissues. We were not able to identify *cis*-eQTLs meeting the selection criteria in the analysed tissues for matrix metalloproteinase 12 (MMP12), ANGPTL4, and BRAP. We found strong evidence supporting the causal effects of the genetically predicted expression level of *ABO* on ASI, PAD, CAD, myocardial infarction, and ischaemic stroke subtypes in the liver, with no confounding by LD as indicated by the HEIDI test (HEIDI P-value ≥ 0.05) (**Supplemental figure 1; ST6**). The associations with PAD and ischemic stroke subtypes in the artery and adipose tissues were also robust and aligned with results from proteome-wide MR analyses. However, we observed discrepancies in several tissue-specific associations, which were opposite to those found in the proteome-wide MR analysis. For example, SMR analysis revealed a positive association between *DUSP13* expression and PAD and CAD in whole blood, which contrasts with the negative effect observed in proteome-wide MR analysis. Similarly, the positive association between zinc finger protein ZPR1 levels and PAD and myocardial infarction was reversed, showing a negative effect when *ZPR1* expression was analysed in the whole blood.

### Triangulation with observational analysis

We triangulated the findings of the 10 proteins with arteriosclerotic/atherosclerotic markers and cardiovascular events using data from the UKB. The observational analysis revealed significant associations between baseline circulating levels of ABO, ANGPTL4, and apolipoprotein B (APOB) with CAD (Hazard ratio (HR): 1.06 per-SD higher ABO; HR: 1.18 per-SD higher ANGPTL4; HR: 1.06 per-SD higher APOB) and myocardial infarction (HR: 1.06 per-SD higher ABO; HR: 1.20 per-SD higher ANGPTL4; HR: 1.06 per-SD higher APOB); baseline levels of fibronectin 1 (FN1) were also significantly associated with CAD (HR: 0.95), and ANGPTL4 levels were significantly associated with PAD (HR: 1.51; **Figure 5**). These results were consistent with the primary MR analysis.

**Figure 5.**
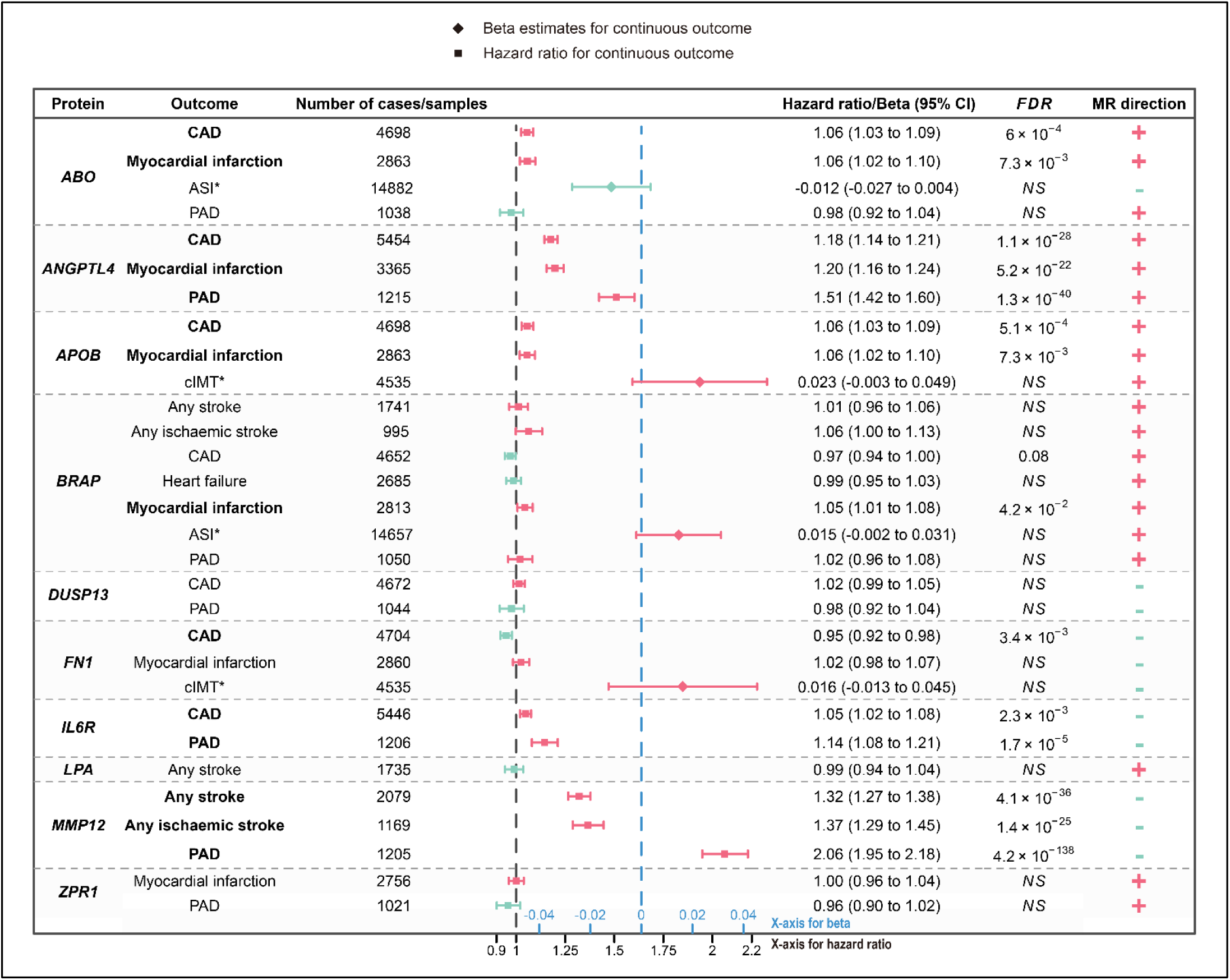
Forest plot showing effects of protein circulating levels on arteriosclerotic/atherosclerotic markers and cardiovascular events from the observational analysis. Hazard ratio estimates were derived from the Cox proportional hazard models for all stroke, ischaemic stroke, CAD, heart failure, myocardial infarction, and PAD (illustrated as rectangles); Beta estimates were derived from the linear regression model for ASI and cIMT (illustrated as diamonds), which were marked with asterisks; FDR-corrected P-values (*P*) were only shown for associations that reached nominal significance; Associations that were FDR significant were highlighted in bold; MR direction column shows the direction of the causal effect from proteome-wide MR analysis; *NS*, not significant; ASI, arterial stiffness index; cIMT, carotid artery intima-media thickness; PAD, peripheral artery disease; CAD, coronary artery disease.

### Mediation effect of protein on cardiovascular events via arteriosclerosis and atherosclerosis

To estimate which proportion of the effect of proteins on cardiovascular events are potentially mediated through arteriosclerotic/atherosclerotic markers, we conducted a mediation analysis. We took the β_EM_ (i.e., effects of proteins on markers) and β_EO_ (i.e., effects of proteins on cardiovascular events) directly from the primary proteome-wide MR analysis. The β_MO_ (i.e., effects of markers on cardiovascular events) were estimated from the bidirectional univariable MR and assessed following rigorous criteria (P_IVW_ < 0.05 and supported by either MR-Egger or WM, and horizontal pleiotropy: P_Egger intercept_ ≥ 0.05; **ST7-8**). Mediator-outcome pathways with potential bidirectional effects were kept to reveal as many potential mediation pathways as possible. As a result, we performed a mediation analysis restricted to 21 protein-arteriosclerosis/atherosclerosis-event pathways (**Figure 6A; ST9**). The most prominent mediation effect was the lipoprotein(a) (LPA) effect on any stroke via carotid plaque (proportion mediated: 92.7%; **Figure 6B**), followed by the DUSP13 effect on CAD via PAD (proportion mediated: 92.7%). All the detailed diagrams showing each mediation pathway were presented **Supplemental Figures 2-4**.

**Figure 6.**
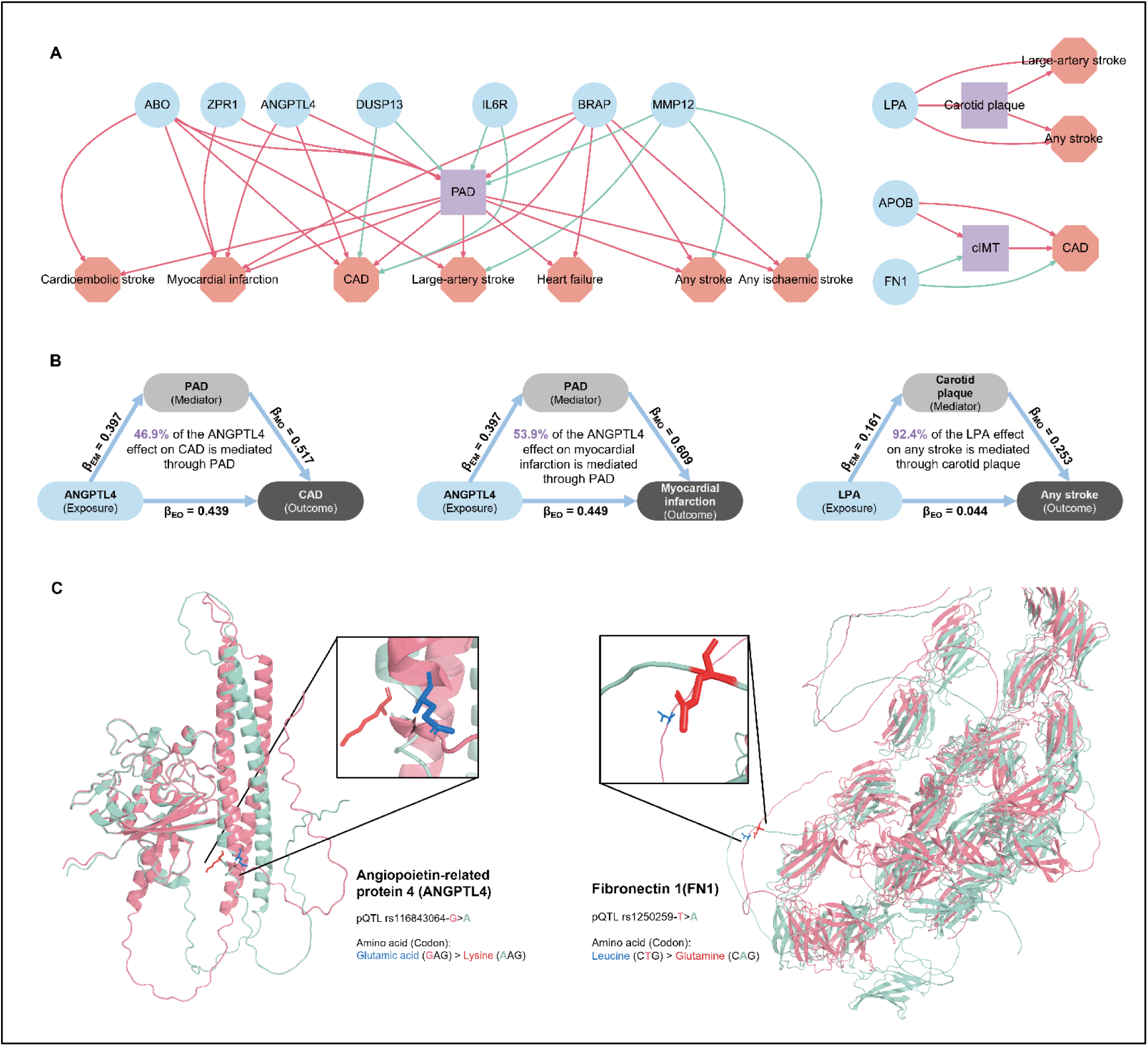
Mediation analyses and 3D structural alterations predicted by AlphaFold 3. **A.** Constructed mediation networks from the mediation analyses. Pathways with positive and negative effects were coloured in red and green, respectively. Circle node: exposure; square node: mediator; octagon node: outcome. **B.** Diagrams showing proportion of protein effect on CAD and stroke outcomes through arteriosclerosis and atherosclerosis. β_EM_, effects of exposure on mediator; β_EO_, effects of exposure on the outcome; β_MO_, effects of mediator on the outcome; cIMT, carotid artery intima-media thickness; PAD, peripheral artery disease; CAD, coronary artery disease. **C.** Left: Predicted structural alterations of ANGPTL4 resulted from the missense variant rs116843064. Right: Predicted structural alterations of FN1 resulted from the missense variant rs1250259. The 3D structure of wild-type protein (reference allele) and mutated protein (alternative allele) were coloured in pink and cyan, respectively. The amino acid encoded by reference codon and alternative codon were highlighted in stick representation and coloured in blue and red, respectively.

### Structural alterations of 7 proteins predicted by AlphaFold3

We screened all proteins associated with arteriosclerotic/atherosclerotic markers (N = 33) and found 7 pQTLs utilized as IVs in MR analyses were either missense variants themselves or in strong LD (*r*^2^ > 0.8) with a missense variant affecting the corresponding protein (**ST10**). **Figure 6C (left)** illustrates the 3D structural alterations of ANGPTL4 resulting from the missense variant rs116843064, which leads to the substitution of glutamic acid (Glu; highlighted in blue) to lysine (Lys; highlighted in red) at position 40. Similarly, **Figure 6C (right)** represents the 3D structural alterations of FN1 resulting from the missense variant rs1250259, which leads to the substitution of leucine for glutamine at position 15. Visualisations of other proteins (including ENTPD6, FRZB, LTBP4, RABEPK, and ZPR1) were presented in **Supplemental Figures 5-9**.

### Investigation of approved and investigational drugs

By integrating information from the Open Targets and DrugBank databases for the 10 proteins prioritized in the previous analyses, we identified several drugs that have been approved or reached phase 4 clinical trials (**ST11-12**). APOB is the target of Mipomersen, an Apo-B 100 mRNA antisense inhibitor used to treat hypercholesterolemia. FN1 is targeted by Ocriplasmin, a proteolytic enzyme indicated for vitreomacular adhesion. IL6R is targeted by several different drugs indicated for diseases including COVID-19, rheumatoid arthritis, giant cell arteritis, systemic sclerosis-associated interstitial lung disease, polyarticular juvenile idiopathic arthritis, systemic juvenile idiopathic arthritis, cytokine release syndrome, and neuromyelitis optica spectrum disorder. LPA has been explored as a target of aminocaproic acid for managing excessive postoperative bleeding. MMP12 is targeted by acetohydroxamic acid, a medication indicated for chronic urea-splitting urinary infections. Additionally, Marimastat, a specific inhibitor of matrix metalloproteinase 12, has been investigated for the treatment of lung and breast cancer.

## Discussion

This study provides novel insights into the shared and distinct molecular pathways that associate arteriosclerosis and atherosclerosis with cardiovascular events. Combining genetic colocalization with MR approaches and further validation using observational epidemiologic data, we identified plasma-based biomarkers of vascular remodelling and CVD progression in different vascular beds. A key finding of this study is the limited overlap of potential causal proteins across the studied markers in different vascular beds, which indicates variation in pathophysiological mechanisms driving atherosclerosis and arteriosclerosis in these regions and highlights the need for vascular-bed-specific treatments. Furthermore, our analysis resulted in the identification of ten candidate proteins (ABO, ANGPTL4, APOB, BRAP, DUSP13, FN1, IL6R, LPA, MMP12, and ZPR1) potentially causally associated with both the arteriosclerotic/atherosclerotic markers and cardiovascular events, validated in two independent datasets (UKB-PPP and deCODE). In addition, integration of proteomic, transcriptomic and observational data confirmed the strength of associations, with proteins including ABO, ANGPTL4, APOB BRAP, and FN1 consistently associating with major clinical endpoints including CAD, myocardial infarction, and stroke. Notably, carotid plaque was a significant mediator that mediated the effect of LPA on stroke, while PAD was a significant mediator linking ABO, ANGPTL4, BRAP, DUSP13, IL6R, ZPR1, and MMP12, to stroke and CAD. By incorporating AlphaFold3 for the prediction of structural alteration, we illustrated the 3D structural alterations of seven proteins induced by the corresponding missense variants, providing additional insights into the underlying biological mechanisms.

Arteriosclerosis, characterised by arterial stiffness, and atherosclerosis, characterised by lipid deposits in arterial walls, have common molecular pathways (including chronic inflammation, oxidative stress and endothelial dysfunction). However, this study identified different protein associations and the heterogeneous effects across vascular beds. For example, PAD and CAD shared many significantly associated protein drivers (e.g., ANGPTL4 and DUSP13), while cIMT and PAD showed few overlapping drivers. Specifically, we observed that proteins such as IL6R and FN1 showed a relationship with cIMT and CAD, while they had no significant association with PAD indicating that the importance of pathways associated with inflammation and extracellular matrix may differ depending on the vascular territory involved. One likely reason is that different vascular markers capture distinct pathophysiological processes: ASI reflects vascular fibrosis and stiffness, CAC indicates calcification, and carotid plaque involves lipid accumulation and inflammation, each representing different stages of vascular disease ^34,35^. Additionally, vascular beds differ in flow dynamics, mechanical stress, and gene expression profiles, shaping region-specific susceptibility to disease. Circulating proteins originate from multiple tissues, including the liver, endothelium, and immune cells, and reflect both systemic and tissue-specific processes modulated by genetic and post-translational mechanisms.

Of the 10 proteins associated with cardiovascular outcomes that we identified in this study, some have established mechanisms linking them to these diseases. APOB is an established atherogenic factor, as it is involved in lipid metabolism, driving the transport of low-density lipoproteins (LDL) that are deposited in the vessel wall leading to plaque development ^36^. ANGPTL4 is associated with lipid metabolism and endothelial function, as it inhibits lipoprotein lipase activity resulting in dyslipidemia and elevation of cardiovascular risk ^37^. IL6R is linked to several inflammatory signalling pathways and its activation leads to chronic inflammation that is pivotal to the development of atherosclerosis and vascular dysfunction ^38^. MMP12 (matrix metalloproteinase 12) is an enzyme that plays a role in extracellular matrix remodelling and has been implicated in destabilization of atherosclerotic plaque, increasing the risk of plaque rupture and subsequent cardiovascular events ^39^. Finally, LPA (lipoprotein[a]) promotes inflammation and increases oxidative stress as well as impairing fibrinolysis, which raises the risk for cardiovascular events like stroke and myocardial infarction ^40^. These mechanistic insights provide further support for the causative associations observed in this study and identify potential targets for therapeutic intervention.

ABO is a pleiotropic locus, meaning its effects may influence multiple traits through diverse and sometimes unrelated pathways. While the associations identified in this study between ABO and vascular traits could theoretically arise through unrelated mechanisms, there are well-established pathways that link the ABO locus to CVD. Non-O blood groups (A, B, AB) are associated with higher levels of von Willebrand factor and factor VIII, which increase thrombosis risk (including risk of ischemic heart disease), as well as potential effects on endothelial function, inflammation, and lipid metabolism ^41,42^. These pathways provide plausible biological explanations for the involvement of ABO in CVD risk and highlight its relevance in vascular pathologies. While in this study, we found that higher circulating ABO levels were associated with an increased risk of PAD, CAD, myocardial infarction, and stroke, whereas they were linked to a lower ASI. Given that arterial stiffness is considered as the direct cause as well as consequence of elevated blood pressure ^43^, our findings align, at least partially, with a previous observational study on the same UKB population, which they observed that non-O group individuals have a higher risk of thromboembolic diseases, but lower risk of hypertension compared to the O group ^44^. This suggests that the ABO blood group may influence vascular pathophysiology through distinct pathways while further investigation is needed to clarify the precise molecular mechanisms.

Integrating AlphaFold3, our study reveals potential molecular mechanisms of action underlying identified causal relationships and further strengthens biological plausibility. We illustrated the structural changes of ANGPTL4 inducing by the missense variant rs116843064 (E40K variant), a pQTL also being utilized as IV in the proteome-MR analysis. This pQTL is associated with reduced circulating levels of ANGPTL4 and lower risk of PAD, CAD, and myocardial infarction. Substantial experimental evidence has demonstrated that E40K variant of ANGPTL4 lowers plasma triglycerides and increases plasma high-density lipoprotein cholesterols by reducing inactivation of lipoprotein lipase through the prevention of extracellular accumulation of ANGPTL4 ^45,46^. Thus, the A allele at rs116843064, leading to the substitution of Glu with Lys, may contribute to a reduced risk of atherosclerosis and CAD.

By identifying key proteins as causal drivers, this research provides a foundation for the development of targeted therapeutic interventions aimed at modulating these pathways to prevent or treat vascular diseases. For instance, proteins like ANGPTL4 and BRAP, associated with both PAD and CAD, could be explored as dual-purpose therapeutic targets to reduce the burden of disease in multiple vascular territories. Moreover, the distinct and shared protein profiles across vascular beds, along with their varied mediation roles, highlight the need for precision medicine approaches that account for the specific pathophysiological context of different CVDs. For instance, previous observational and MR studies have established elevated LPA levels as a risk factor for large-artery stroke ^47,48^. Our study further revealed that this effect is primarily mediated through the formation of carotid plaque, suggesting LPA as a potential therapeutic target for carotid atherosclerosis. Targeting LPA may offer a strategy to reduce the risk of large-artery stroke by mitigating plaque development and its associated vascular complications. In addition, the intersection with existing and investigational drug-target databases identified many of the prioritized proteins (e.g., APOB, IL6R, LPA, and MMP12) as already druggable with existing or investigational compounds. For example, Mipomersen’s targeting of APOB for hypercholesterolemia demonstrates its clinical relevance for lipid-associated cardiovascular risk ^49^. Notably, the ongoing trials with IL6R inhibitors in inflammatory diseases may provide new opportunities for repurposing these drugs already known to benefit CVD.

### Strengths and Limitations

A great strength of this study is the integration of large-scale proteomic, transcriptomic, and genetic data to illuminate shared causal pathways across arteriosclerosis, atherosclerosis, and diverse vascular phenotypes. Bayesian colocalization with additional stringent bidirectional MR analyses, minimize biases from confounding and reverse causation allowing for even stronger data supporting our findings. Specifically, we performed Bayesian colocalization to pinpoint a shared genetic region with strong evidence of sharing the same causal variant for a given protein-trait pair prior to selecting IVs for causal appraisal, which help reducing the risk of confounding by LD and horizontal pleiotropy ^50^. Second, to the best of our knowledge, this is the first study to examine the potential mediating role of arteriosclerosis and atherosclerosis in the association between plasma proteins and cardiovascular events. This helps clarify the specific mechanisms by which the plasma protein contributes to CVD risk, enhancing further causal interpretation. Nonetheless, there are a few important limitations to discuss. First, the study used data from predominantly individuals of European ancestry, restricting the generalizability of its findings to other populations. Second, MR analysis relies on several key assumptions, one of which is the absence of horizontal pleiotropy (i.e., exclusion restriction). Violation of this assumption could introduce bias into the results, and while robust MR methods can mitigate some of the effects, they cannot completely exclude the possibility of horizontal pleiotropy ^51^. In addition, casual estimates derived from MR reflect the lifelong effect of an exposure on an outcome, rather than the short-term or modifiable effect derived from conventional observational analyses or clinical trials. Therefore, these estimates should be interpreted with caution. Third, several protein-trait associations demonstrated unconcordant results in SMR utilizing tissue-specific eQTLs comparing the primary MR analysis utilizing plasma pQTLs. Although this does not invalidate our findings, one possible interpretation might be that the observed causal effects may depend on post-transcriptional regulation or post-translational modifications, such as alternative splicing and phosphorylation. Current available datasets have limited power in identifying significant quantitative trait loci associated with alternative splicing (sQTLs) and isoform-specific QTL (isQTLs) associated with the expression of specific isoforms, hindering further verification. Fourth, validation with observational data entails the potential for residual confounding. The absence of pre-specified ischemic stroke subtypes in the UKB data limits the ability to further triangulate potential causal proteins in relation to specific ischemic stroke subtypes, which exhibited distinct arterial pathologies. Last, Participants included in the CAD GWAS may have also contributed to the UKB-PPP dataset, potentially leading to attenuated estimates of causal effects due to sample overlap.

## Conclusions

In summary, our results provide valuable insights into the complex relationships between circulating plasma proteins and subclinical markers of atherosclerosis and arteriosclerosis across different vascular beds. By employing Bayesian colocalization analysis, followed by a two-sample bidirectional MR approach, our study pinpoints ten plasma proteins with causal evidence linking atherosclerotic and arteriosclerotic pathophysiology to CVD outcomes. Our mediation analysis underscores the critical role of atherosclerosis and arteriosclerosis in modulating these associations, highlighting potential therapeutic targets for precision cardiovascular medicine. Notably, by innovatively incorporating AlphaFold3 into our pipeline, we translate the missense pQTL for corresponding causal proteins from MR analyses into structural alterations, offering more insights into the underlying pathogenicity and molecular mechanisms. While these results offer promising avenues for biomarker-driven risk stratification and potential dual-purpose therapeutic interventions, they also underscore the need for validation in diverse populations and a deeper investigation into the underlying biological mechanisms.

## Data Availability

The UKB-PPP data are available at http://ukb-ppp.gwas.eu, and the deCODE genetics proteomics data can be accessed at https://www.decode.com/summarydata/. GWAS summary statistics for ASI and CAC are available from the GWAS Catalog (https://www.ebi.ac.uk/gwas/) under accession numbers GCST008403 and GCST90278456, respectively. Data for cIMT and carotid plaque were obtained from dbGaP under CHARGE study accession number phs000930.v6.p1, and for PAD under accession number phs001672.v2.p1. Stroke and its subtypes are available via the GWAS Catalog (GCST90104534 to GCST90104563). Summary statistics CAD, myocardial infarction, and heart failure are also accessible through the GWAS Catalog under accession numbers GCST005194, GCST011365, and GCST009541, respectively.

## Ethics declarations

Our study is based entirely on publicly available, summary-level GWAS data. No new individual-level data were collected or analyzed. All original studies from which the data were obtained had received the appropriate ethical approvals.

