## Supplemental Figures for "Shared proteomic landscape between arteriosclerosis and cardiovascular endpoints: a Mendelian randomization and observational study integrating AlphaFold3 for structural prediction" (from another version of this preprint: version 1, 2025-05-16)

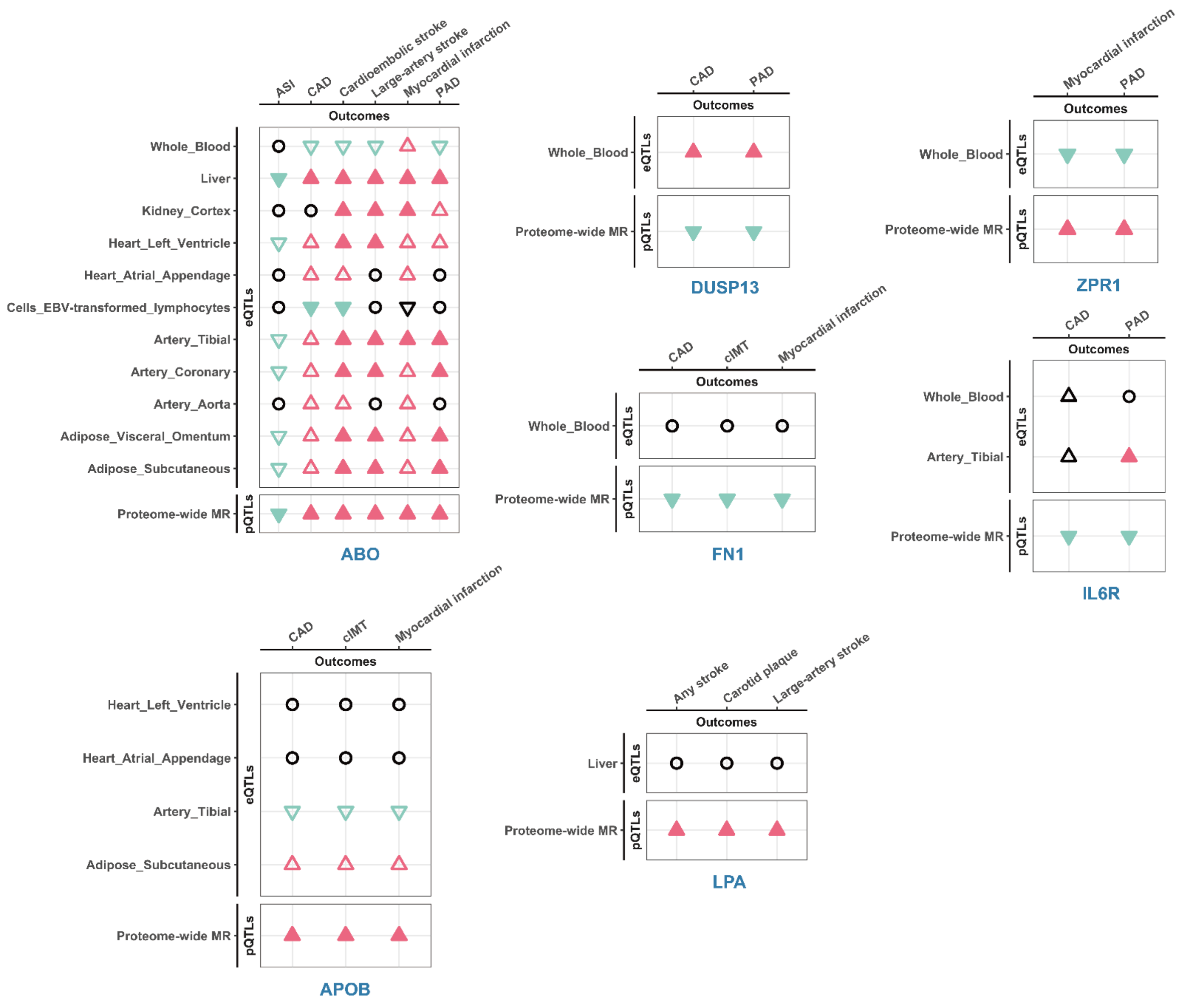


**Supplemental figure 1. Results from transcriptome-wide analysis applying SMR and HEIDI.** The Y-axis displays the names of tissues in which gene expression levels were proxied by eQTLs as the exposure; the lower panels labels as pQTLs show the results from the previous proteome-wide MR analysis. The X-axis displays the names of outcomes (arteriosclerotic/atherosclerotic markers and cardiovascular events). Triangle: nominal significant; Coloured triangle: FDR significant; Filled triangle: pass HEIDI test; Hollow triangle: does not pass HEIDI test; Circle: not significant; The direction of the triangle represents the direction of the association. ASI, arterial stiffness index; cIMT, carotid artery intima-media thickness; PAD, peripheral artery disease; CAD, coronary artery disease.

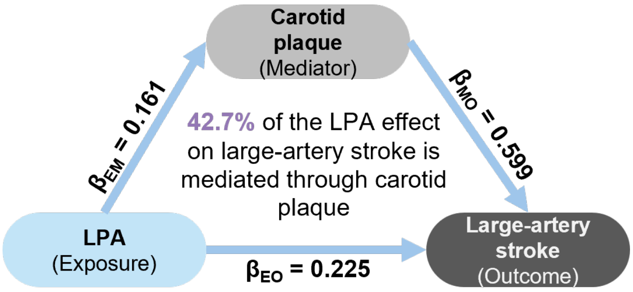


**Supplemental figure 2. Diagrams showing proportion of protein effect on CAD and stroke outcomes through carotid plaque.**

$\beta_{EM}$, effects of exposure on mediator; $\beta_{EO}$, effects of exposure on the outcome; $\beta_{MO}$, effects of mediator on the outcome.

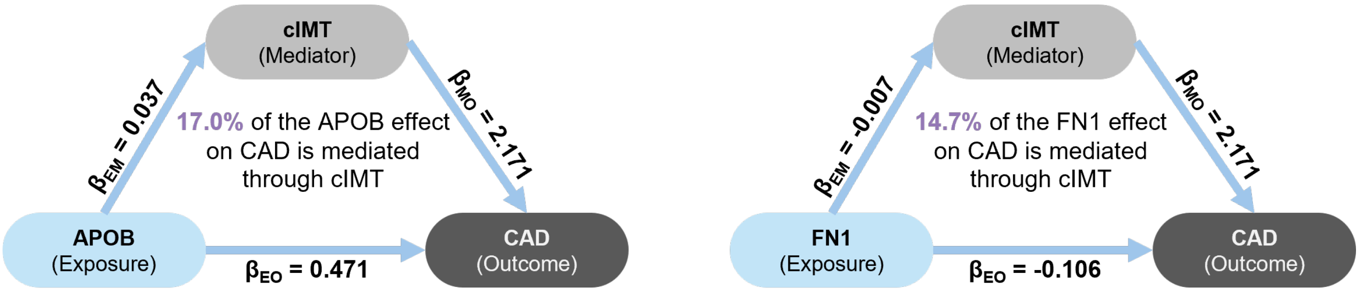


**Supplemental figure 3. Diagrams showing proportion of protein effect on CAD and stroke outcomes through cIMT.** $\beta_{EM}$, effects of exposure on mediator; $\beta_{EO}$, effects of exposure on the outcome; $\beta_{MO}$, effects of mediator on the outcome; cIMT, carotid artery intima-media thickness; CAD, coronary artery disease.

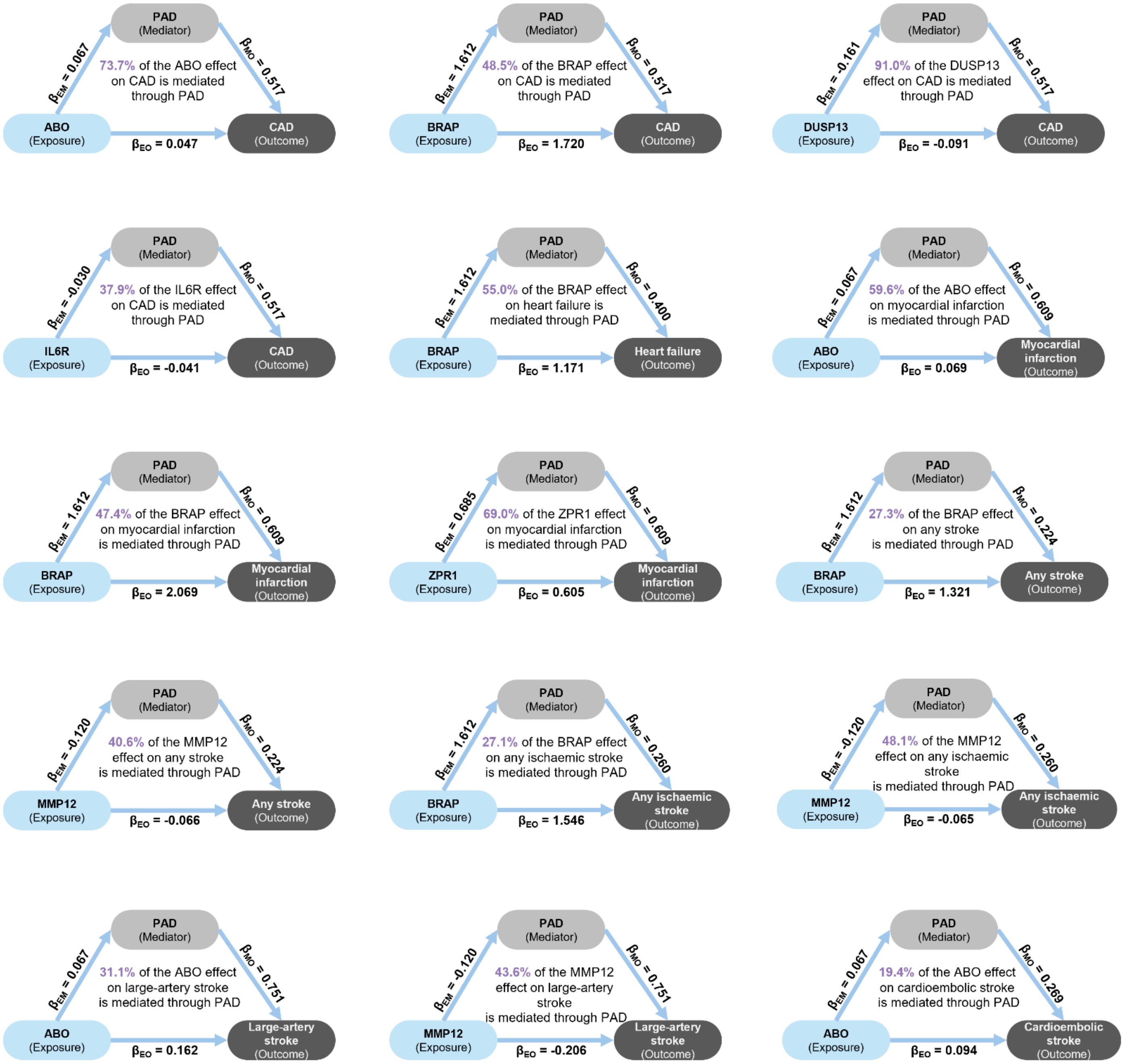


**Supplemental figure 4. Diagrams showing proportion of protein effect on CAD and stroke outcomes through PAD.** $\beta_{EM}$, effects of exposure on mediator; $\beta_{EO}$, effects of exposure on the outcome; $\beta_{MO}$, effects of mediator on the outcome; PAD, peripheral artery disease; CAD, coronary artery disease.

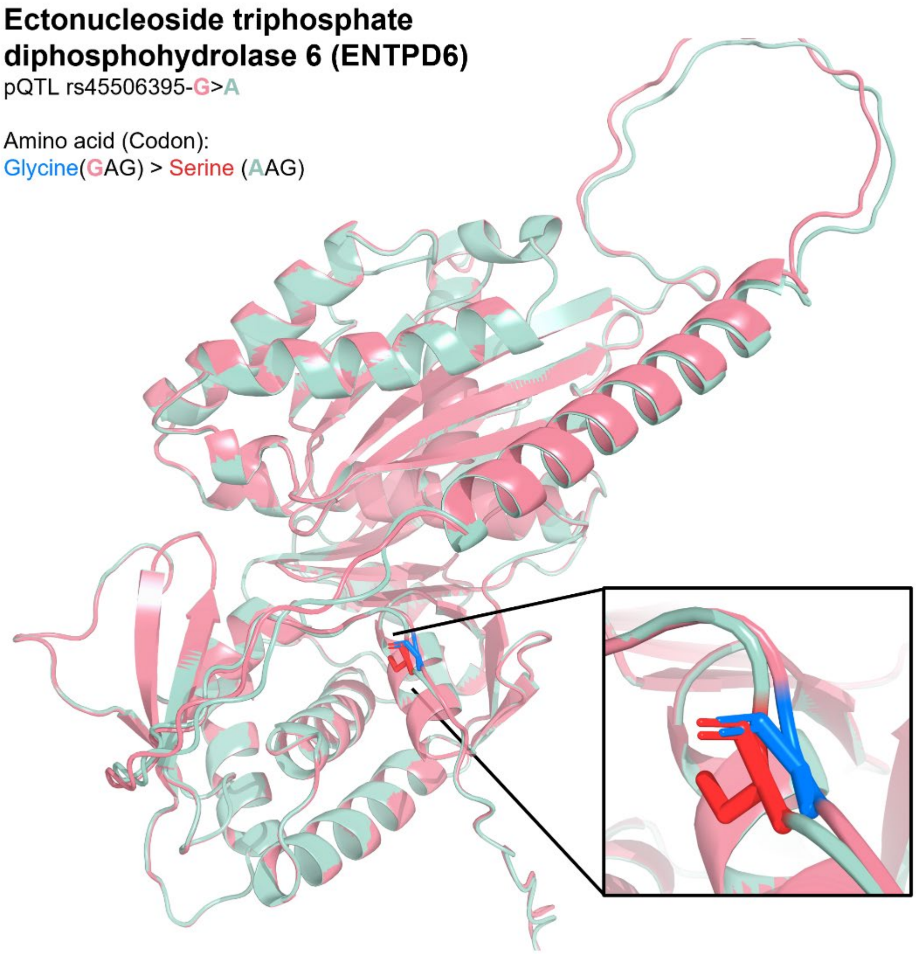


**Supplemental figure 5. Predicted structural alterations of ENTPD6 resulted from the missense variant rs45506395.** The 3D structure of wild-type protein (reference allele) and mutated protein (alternative allele) were coloured in pink and cyan, respectively. The amino acid encoded by reference codon and alternative codon were highlighted in stick representation and coloured in blue and red, respectively.

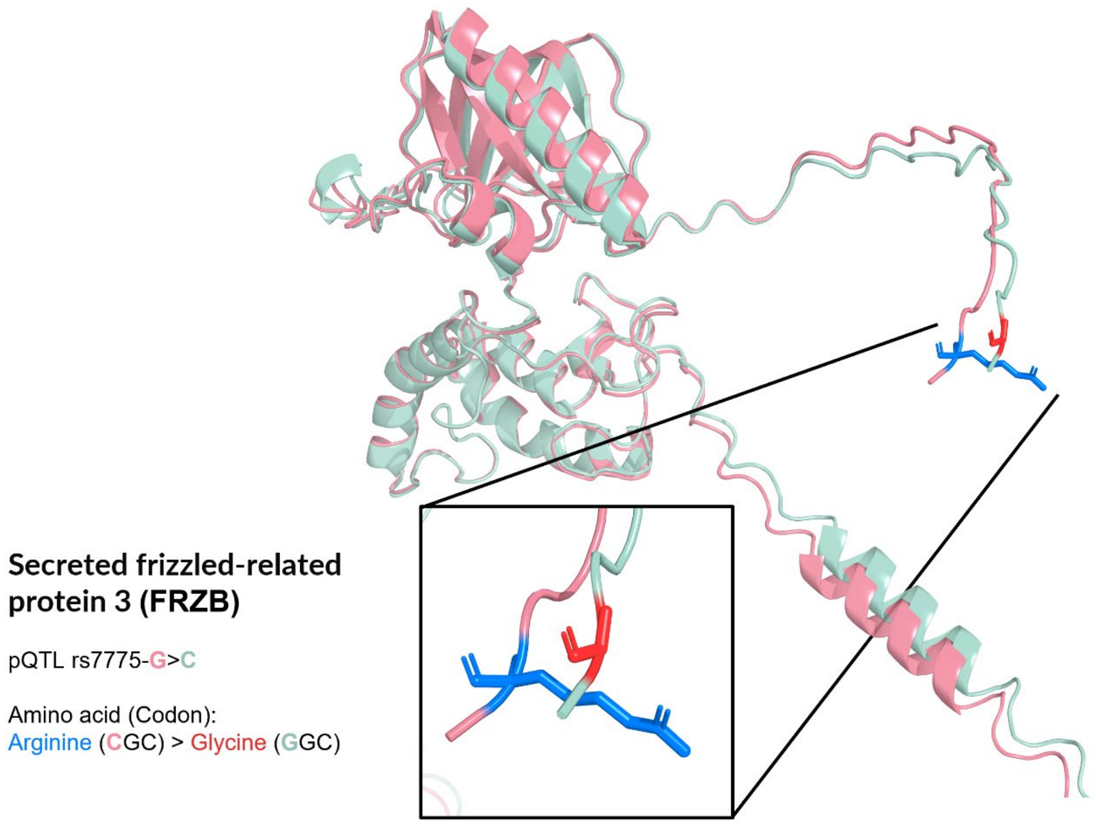


**Supplemental figure 6. Predicted structural alterations of FRZB resulted from the missense variant rs7775.** The 3D structure of wild-type protein (reference allele) and mutated protein (alternative allele) were coloured in pink and cyan, respectively. The amino acid encoded by reference codon and alternative codon were highlighted in stick representation and coloured in blue and red, respectively.

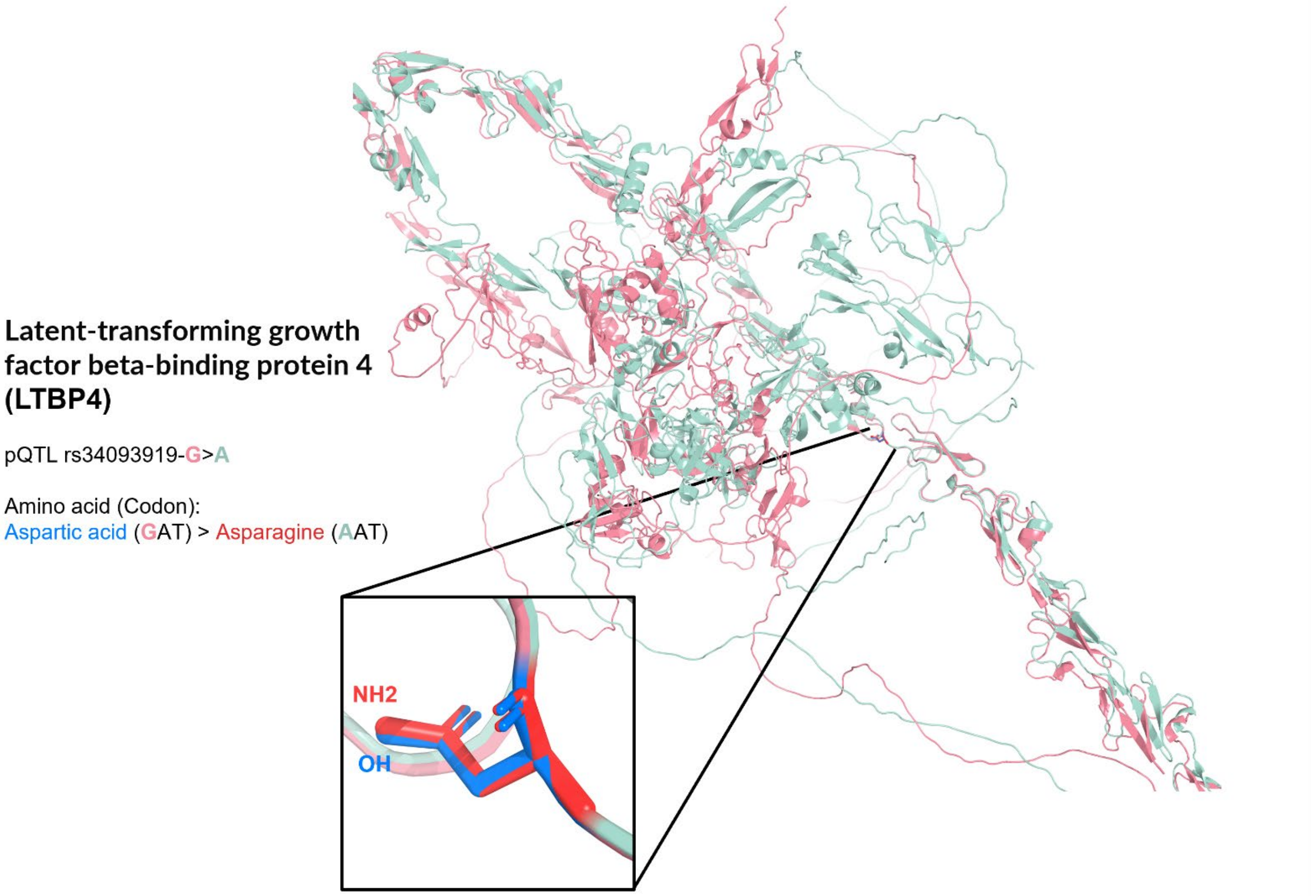


**Supplemental figure 7. Predicted structural alterations of LTBP4 resulted from the missense variant rs34093919.** The 3D structure of wild-type protein (reference allele) and mutated protein (alternative allele) were coloured in pink and cyan, respectively. The amino acid encoded by reference codon and alternative codon were highlighted in stick representation and coloured in blue and red, respectively.

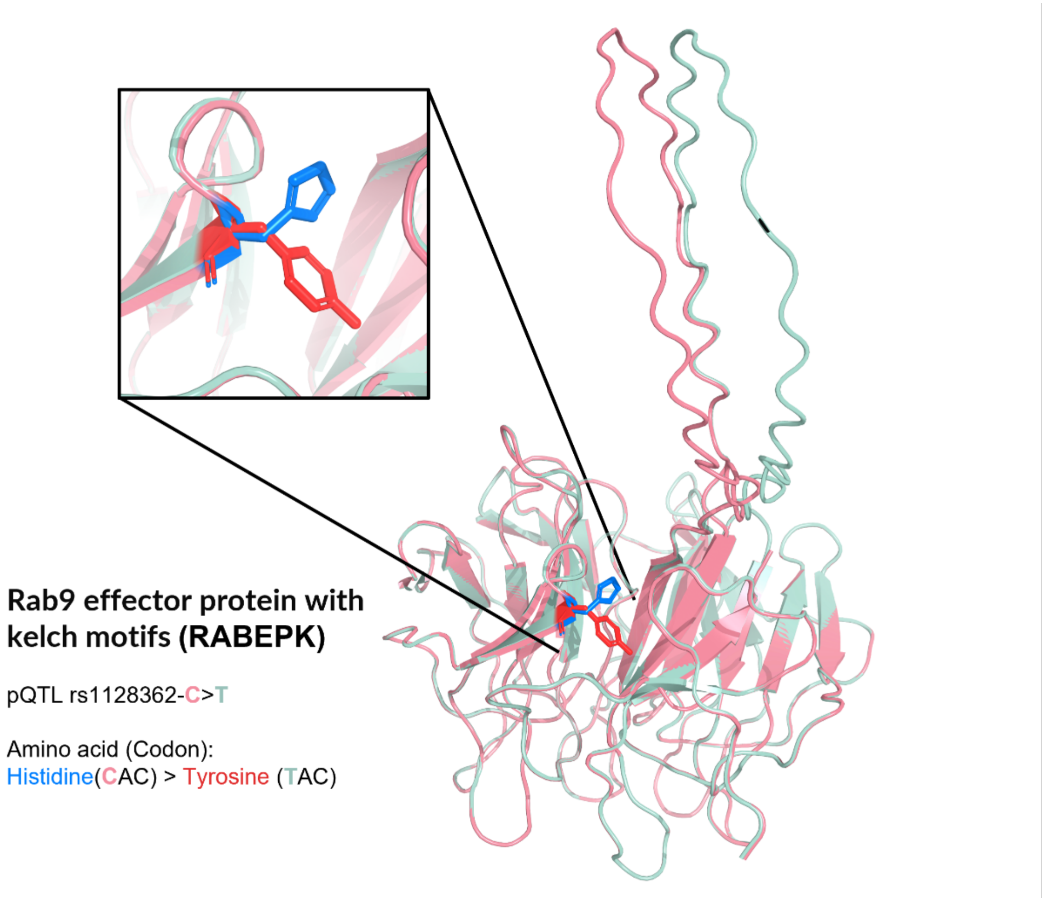


**Supplemental figure 8. Predicted structural alterations of RABEPK resulted from the missense variant rs1128362.** The 3D structure of wild-type protein (reference allele) and mutated protein (alternative allele) were coloured in pink and cyan, respectively. The amino acid encoded by reference codon and alternative codon were highlighted in stick representation and coloured in blue and red, respectively.

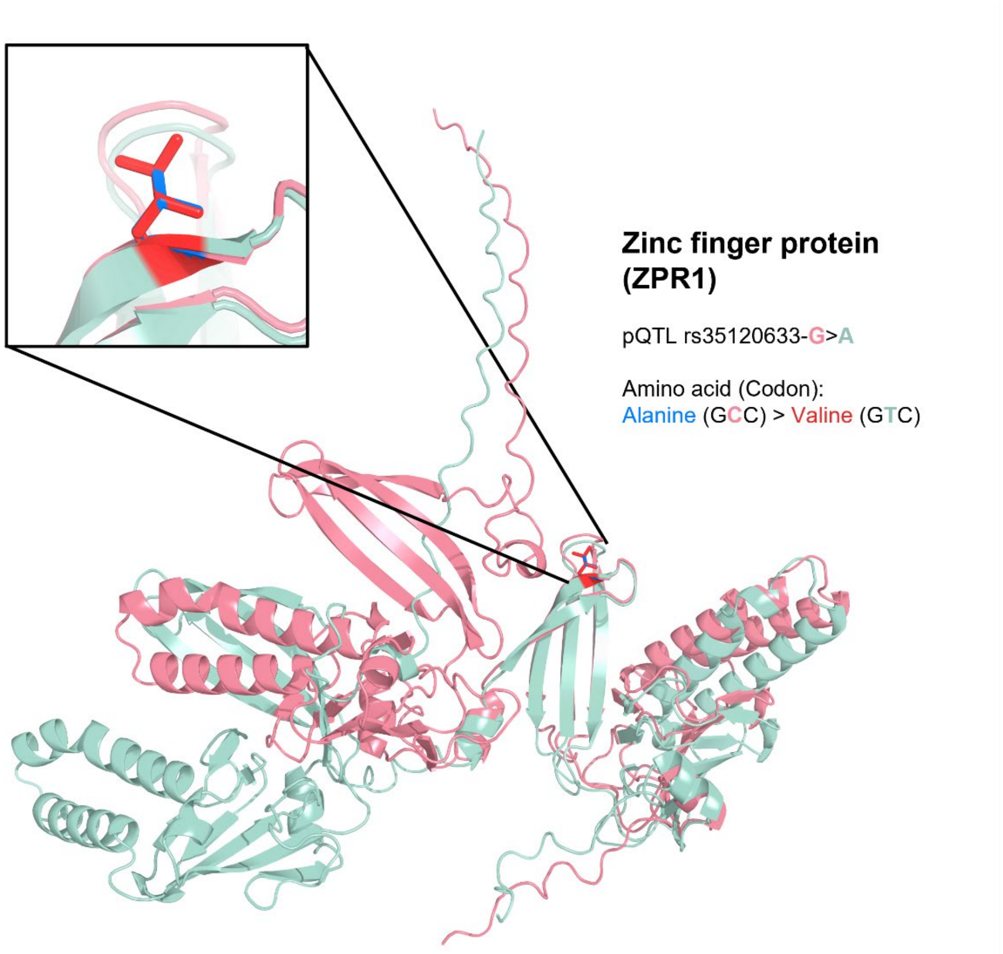


**Supplemental figure 9. Predicted structural alterations of ZPR1 resulted from the missense variant rs35120633.** The 3D structure of wild-type protein (reference allele) and mutated protein (alternative allele) were coloured in pink and cyan, respectively. The amino acid encoded by reference codon and alternative codon were highlighted in stick representation and coloured in blue and red, respectively.
